# Paediatric Personalized Research Network Switzerland (SwissPedHealth): A Joint Paediatric National Data Stream

**DOI:** 10.1101/2024.07.24.24310922

**Authors:** Rebeca Mozun, Fabiën N. Belle, Andrea Agostini, Matthias R. Baumgartner, Jacques Fellay, Christopher B. Forrest, D. Sean Froese, Eric Giannoni, Sandra Goetze, Katrin Hofmann, Philipp Latzin, Roger Lauener, Aurélie Martin Necker, Kelly E. Ormond, Jana Pachlopnik Schmid, Patrick G. A. Pedrioli, Klara M. Posfay-Barbe, Anita Rauch, Sven Schulzke, Martin Stocker, Ben D. Spycher, Effy Vayena, Tatjana Welzel, Nicola Zamboni, Julia E. Vogt, Luregn J. Schlapbach, Julia A. Bielicki, Claudia E. Kuehni, SwissPedHealth consortium

## Abstract

**Introduction:** Children represent a large and vulnerable patient group. However, the evidence-base for most paediatric diagnostic and therapeutic procedures remains limited or is often inferred from adults. There is urgency to improve paediatric health care provision based on real-world evidence generation. The digital transformation is a unique opportunity to shape a data-driven, agile, learning health care system and deliver more efficient and personalized care to children and their families. The goal of SwissPedHealth is to build a sustainable and scalable infrastructure to make routine clinical data from paediatric hospitals in Switzerland interoperable, standardized, quality-controlled, and ready for observational research, quality assurance, trials, and health-policy creation. This paper describes the design, aims, and current achievements of SwissPedHealth.

**Methods and analysis:** SwissPedHealth started in September 2022 as one of four National Data Streams co-funded by the Swiss Personalized Health Network (SPHN) and the Personalized Health and Related Technologies (PHRT). SwissPedHealth develops modular governance and regulatory strategies, and harnesses SPHN automatization procedures, in collaboration with clinical data warehouses, the Data Coordination Center, Biomedical Information Technology Network, and other SPHN institutions and funded projects. The SwissPedHealth consortium is led by a multi-site, multi-disciplinary Steering Committee incorporating patient and family representatives. The data stream contains work packages focusing on: 1) governance and implementation of standardized data collection, 2) nested projects to test the feasibility of the data stream, 3) a lighthouse project that enriches the data stream by integrating multi-omics data, aiming to improve diagnoses of rare diseases, and 4) engagement with families through patient and public involvement activities and bioethics interviews.

**Ethics and dissemination:** The health database regulation of SwissPedHealth was approved by the ethics committee (AO_2022-00018). Research findings will be disseminated through national and international conferences, publications in peer-reviewed journals and in lay language via online media and podcasts.

**ARTICLE SUMMARY:**

⍰ The paediatric national data stream SwissPedHealth focuses on routine clinical data from children at Swiss University Children’s Hospitals, spans disciplines and is built in a scalable and modular way in terms of governance, data infrastructure, and patient and public involvement, to enable a gradual increase in coverage of the Swiss child population.
⍰ SwissPedHealth strives to increase readiness for quality improvement, research, and personalized paediatric health care.
⍰ SwissPedHealth’s infrastructure aligns with the national frameworks, safeguarding data security and adhering to a standard interoperability framework based on the Resource Description Framework (RDF).
⍰ SwissPedHealth seeks to explore integration of data from external sources such as federal statistics, cohorts, and registries, which have their own governance and data formats.
• SwissPedHealth investigates the use of multi-omics workflows to improve diagnosis of rare diseases in children with life-threatening phenotypes

## INTRODUCTION

In Switzerland, over 85,000 children are born annually, contributing to a current population of 1.7 million inhabitants aged under 20 years.^1^ Every year, over 100,000 children require admission as inpatients to hospitals. Many more are seen in outpatient clinics. Children represent a vulnerable population, with unique developmental physiology, dependence on adults for care-seeking, and disease epidemiology. Adverse effects related to diseases, treatments, environmental exposures, and lifestyle during early childhood will affect children themselves, their families, and future offspring for many decades, markedly increasing the associated direct health care and other indirect costs for society.^2–5^ The evidence to guide best paediatric health care remains sparse. At present, the evidence-base for many diagnostic and therapeutic procedures is either absent, of low-quality (often data from small cohorts), or inferred from adults. Paediatric evidence generation often suffers from a long lag time from results to implementation. While these features apply to the paediatric population in general, the limitations are aggravated in Switzerland due to the fragmentation of its health care system with regional isolated data repositories. In addition, multiple paediatric registries coexist in Switzerland, and these lack interoperability. Overall, the present state of paediatrics thus lacks preparedness to appropriately develop, evaluate, or adapt to the rapidly increasing possibilities for intelligent decision-support tools which could enhance management of paediatric patients.

Over the past decades, progress in social and preventive health and new therapies have reduced the morbidity and mortality caused by common conditions such as infections, cancer, or trauma. Consequently, modern paediatrics in high-income countries like Switzerland deals with a conundrum of rare diseases which present as unique and often life-threatening phenotypes and require highly personalized approaches.^6–10^ In addition, common problems such as obesity-related morbidity and chronic respiratory conditions frequently result in lifelong trajectories reaching out well into adulthood.^11,12^ Furthermore, although patients and parents have intimate knowledge of their disease and needs, they remain poorly integrated into health care planning and research. The demand to continuously improve paediatric data for research and improved care requires better digitalization, harmonization, and automated extraction of routine clinical data. This represents a unique opportunity to break silos and to shape a data-driven, agile, learning health care system that will deliver better, more efficient, and more personalized care to children and their families. SwissPedData, an earlier Swiss initiative, defined an agreed set of common data elements from routine health care records to gather standardized and high-quality information on key aspects of interest to clinicians and researchers for paediatric clinical care. ^13,14^ This paediatric core dataset has the potential to be enriched with patient-reported outcomes, population-based and administrative information, specialized disease registries, and high-density data from advanced clinical diagnostics and research projects such as omics technologies. As part of a national call for data streams (multidisciplinary consortia research platforms), we designed the Paediatric Personalized Research Network Switzerland (SwissPedHealth).

SwissPedHealth’s main goal is to implement routine clinical data extraction pipelines in paediatric hospitals, make this core dataset available for research, benchmarking, and improvement of quality of care, thereby implementing the infrastructure towards a learning national paediatric health system. This paper describes the design, structure, aims, and current achievements of the national data stream SwissPedHealth.

## METHODS AND ANALYSIS

### SwissPedHealth

SwissPedHealth (https://www.swisspedhealth.ch/) is a paediatric national data stream (NDS) that started in September 2022 aiming to make health-related personal data from routine clinical practice collected from hospitals in Switzerland interoperable, standardized, quality-controlled, and ready for observational research, clinical trials, personalized medicine, health-policy research, and clinical audits. Several quality improvement and research projects will test the infrastructure for routine clinical data collection and enrich the data stream.

The SwissPedHealth data stream is embedded in an ecosystem supported by two funding bodies, the Swiss Personalized Health Network (SPHN, https://sphn.ch/) and the Strategic Focal Area “Personalized Health and Related Technologies (PHRT)” of the ETH Domain (Swiss Federal Institutes of Technology, https://www.sfa-phrt.ch/). SPHN and PHRT aim to build a Swiss personalized health ecosystem, improve research and innovation in health care by developing industrialized infrastructures for efficient secondary use of electronic health record data across hospitals, research institutions, biobanks, and registries in Switzerland and enhance the quality of the collected data.^15^ The Data Coordination Center (https://sphn.ch/network/data-coordination-center/) is managed by the personalized health informatics group of the Swiss Institute of Bioinformatics (https://www.sib.swiss/). The Data Coordination Center promotes, designs, implements, and coordinates efforts around data semantics and standards across Swiss hospitals and other data providers. SIB provides legal support and coordination within the SPHN framework. BioMedIT (https://www.biomedit.ch/) acts as a national secure computing network for health-related data.

The SwissPedHealth consortium includes clinical partners from all five Swiss paediatric university hospitals (Universitäts-Kinderspital beider Basel – UKBB, Inselspital Bern, Hôpitaux universitaires de Genève – HUG, Centre hospitalier universitaire Vaudois – CHUV, and University Children’s Hospital Zurich) and two non-academic hospitals (Luzerner Kantonsspital and Ostschweizer Kinderspital); three research institutions, the Federal Institutes of Technology ETH Zurich (*Eidgenössische Technische Hochschule*), and EPFL (*École Polytechnique Fédérale de Lausanne*), and the Institute of Social and Preventive Medicine (ISPM, University of Bern); and patient and public representatives. The consortium unites expertise in clinical paediatrics and paediatric research, data science, omics, machine learning, patient and public involvement, and public health. SwissPedHealth builds on previous projects including SwissPedData,^13,14^ and the Swiss Research Network of Clinical Paediatric Hubs (SwissPedNet, https://www.swisspednet.ch/home)

### SwissPedHealth governance structure and regulatory framework

The governance structure of SwissPedHealth consists of a steering level with an overarching Steering Committee, an Infrastructure Executive Board, and a Scientific Advisory Board; a management level comprising the Executive Office (Executive Board and Management Office); and an operational level with five work packages covering the data integration and research programs (**Fig. 1**). The regulatory framework of SwissPedHealth builds on existing collaboration agreements through SwissPedNet as well as template documents from SPHN. SwissPedHealth uses a three-tiered structure, namely Tier 1) SwissPedNet Service Level Agreements, Tier 2) SwissPedHealth Infrastructure Consortium Agreement, and Tier 3) project specific Data Project Consortium Agreements. Tier 1 regulates the data flow from participating Children’s Hospital to their Clinical Data Warehouses (CDWs). Tier 2 issues guidance and templates to access clinical data through SwissPedHealth, including General Terms and Conditions for data projects, Data Transfer and Use Agreements, and Data Project Consortium Agreements templates. Tier 3 focuses on project specific governance aspects, and details project specific Data Project Consortium Agreements elements including project aims, sponsorship, project partners, data needs, and timelines. Principles of data governance and regulatory compliance respect primary ownership of routine data by the hospitals with their respective CDWs, and research-only data by the respective research teams. Requests to access data is be governed accordingly at the level of the NDS Data Access Request Committee of the NDS Infrastructure Executive Board. The Infrastructure Consortium Agreement has been approved by the seven participating children’s hospitals and all three BioMedIT nodes.

**Figure 1:**
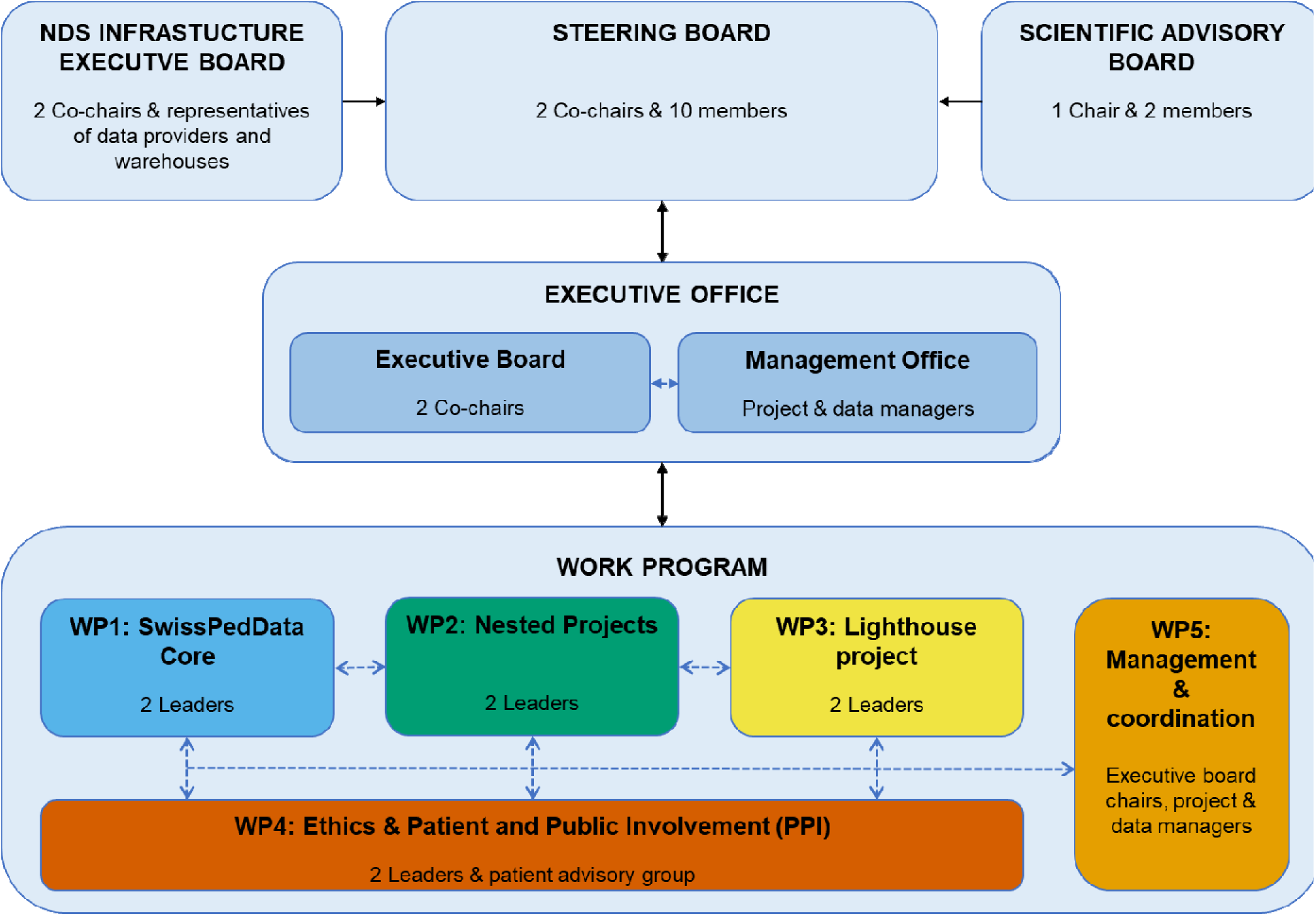
SwissPedHealth governance

### Central data management

#### Data flow, FAIRification, data quality assessment

Routine clinical data is extracted from the electronic data capture systems and other source systems and loaded into the CDWs of each children’s hospital from where it can be transferred to the NDS. Data is harmonized according to the SPHN ontology in each CDW. Data that forms part of the NDS is planned to be indexed as part of the data FAIRification process to make data Findable, Accessible, Interoperable, and Reusable.^16,17^ These FAIR principles lay the basis of common practices among hospitals and facilitate the further use of routine clinical data for research. The data from the different hospital source systems are transformed and mapped to the SwissPedHealth project-specific schema in the CDWs. The project-specific schema extends the SPHN schema to cover the needs of the SwissPedHealth NDS. To achieve an interoperable data representation, that is coherent and semantically understandable both for humans and machines, SPHN employs concepts which incorporate existing ontologies and terminologies such as Systematic Nomenclature of Medicine – Clinical Terms (SNOMED CT, https://www.snomed.org).^15,18^ Concepts are based on the Resource Description Framework (RDF, https://www.w3.org/RDF/). CDWs use the SPHN Connector tool (https://git.dcc.sib.swiss/sphn-semantic-framework/sphn-connector) to pseudonymize and de-identify data, and to transform it to RDF format compliant with the SPHN schema. RDF data integrity checks and data queries can be done using Shapes Constraint Language (SHACL) and SPARQL, an RDF Query Language. Data is transferred from CDWs to the secure project space (so called “B-space”) for the SwissPedHealth NDS at the sciCORE BioMedIT node in Basel via the Secure Encryption and Transfer Tool (SETT) tool (https://gitlab.com/biomedit/sett) (**Fig. 2)**. Data transfers from CDWs to the NDS B-space are planned to occur recurrently.

**Figure 2:**
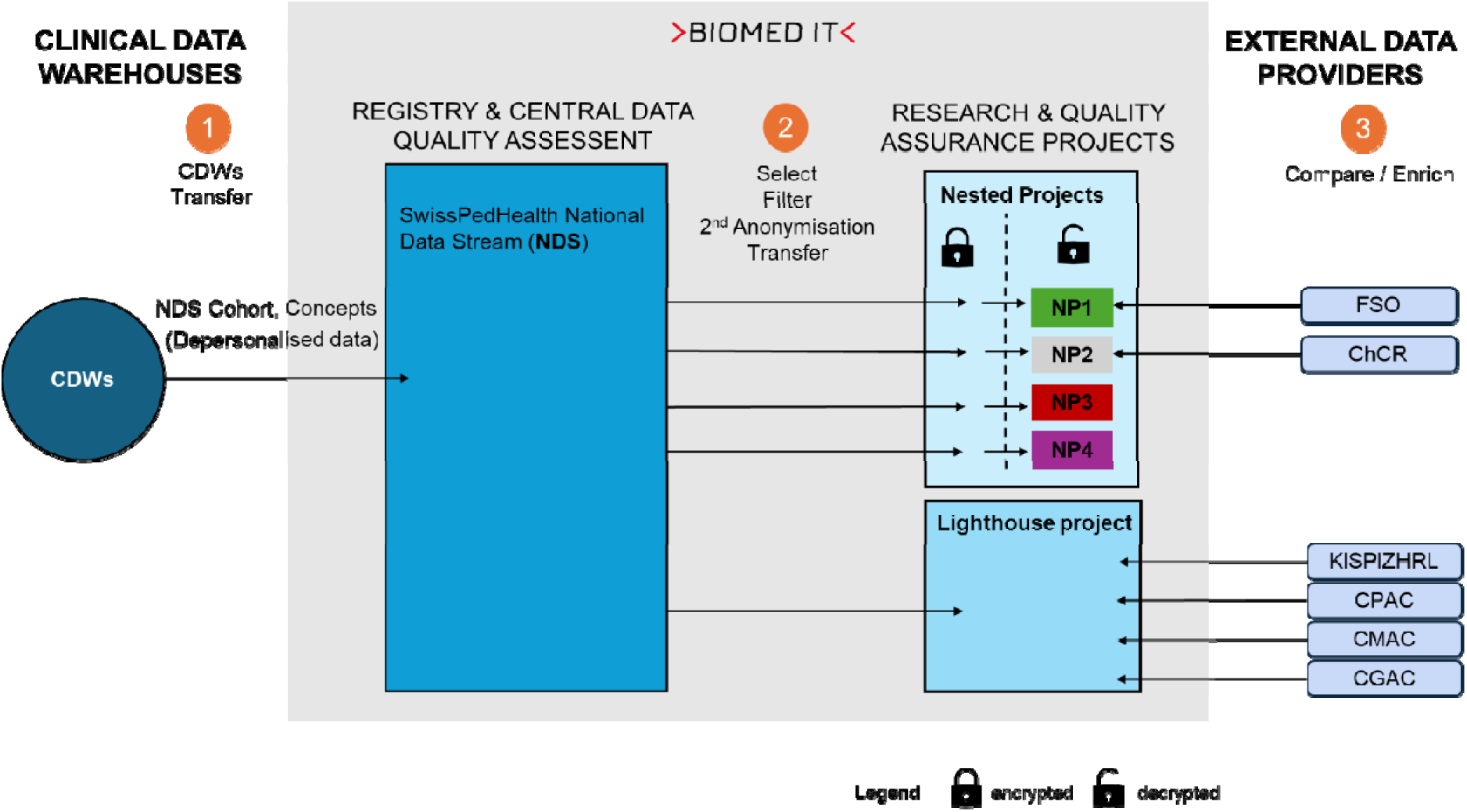
SwissPedHealth data flow **Abbreviations:** CDWs = clinical data warehouses; LH = Lighthouse project; NDS = National Data Stream; NP1 = SwissPedGrowth, NP2= SwissPedCancer, NP3 = SwissPedLung, NP4 = SwissPedAntibio; FSO = Federal Statistical Office; ChCR = Swiss childhood cancer registry; CPAC = Clinical Proteomics Analysis Centre; CMAC = Metabolomics Analysis Centre; CGAC = Clinical Genomic Analysis Centre; KISPIZHRL = University Children’s Hospital (Kispi) Zurich Research Lab. Numbers indicate:

1) Data flow from CDWs to NDS. Data transfer requests triggered by NDS B-space to CDWs, transfer via the Secure Encryption and Transfer Tool (SETT) tool.
2) Data flow from NDS to project specific spaces. Data transfer triggered by nested projects B-space or lighthouse B-space to NDS B-space.
3) Data flow from external data providers to project specific spaces.

The NDS B-space serves as a data repository, where the central data managers perform data quality assessment and remediation as needed, data filtering, and transfer to separate project specific B-spaces for research or quality improvement projects (called “workspaces”). Currently, SwissPedHealth holds two separate workspaces for data analyses: one for the nested projects and another for the lighthouse project. The access to these separate project-specific workspaces is coordinated by permission managers and restricted to permitted users. Users must perform the mandatory data protection and IT security awareness training of SPHN and accept the sciCORE Terms of Use. Secure and encrypted modalities provided by the SPHN infrastructure are used for all data transfers between CDWs and BioMedIT, and between external data providers and BioMedIT.

#### The nested projects

SwissPedHealth includes four nested projects which serve as use cases to address relevant questions in child health, focusing on 1) anthropometrics – SwissPedGrowth, 2) childhood cancer – SwissPedCancer, 3) paediatric lung function and respiratory diseases – SwissPedLung, and 4) antibiotic utilization – SwissPedAntibio (**Table 1**). These projects showcase the feasibility and usefulness of the NDS, using selected key concepts from the SwissPedData core dataset from a large number of children admitted to Swiss hospitals or visiting outpatient clinics, and at the same time serve as data quality assurance.^19,20^ Health-related personal data obtained in clinical routine, such as demographic and treatment data as well as medical history captured in electronic health records of patients from participating children’s hospitals will be extracted for the years 2017-2023. In the future, regular ongoing data transfers are planned. Transferred data will include SPHN concepts (https://www.biomedit.ch/rdf/sphn-schema/sphn/2024/1), such as diagnostic codes (SNOMED CT, ICD-10, ICD-O3), age, height, and weight, Logical Observation Identifiers, Names and Codes (LOINC) for lab results, and Anatomical Therapeutic Chemical Classification System (ATC) codes for drugs.

**Table 1:**
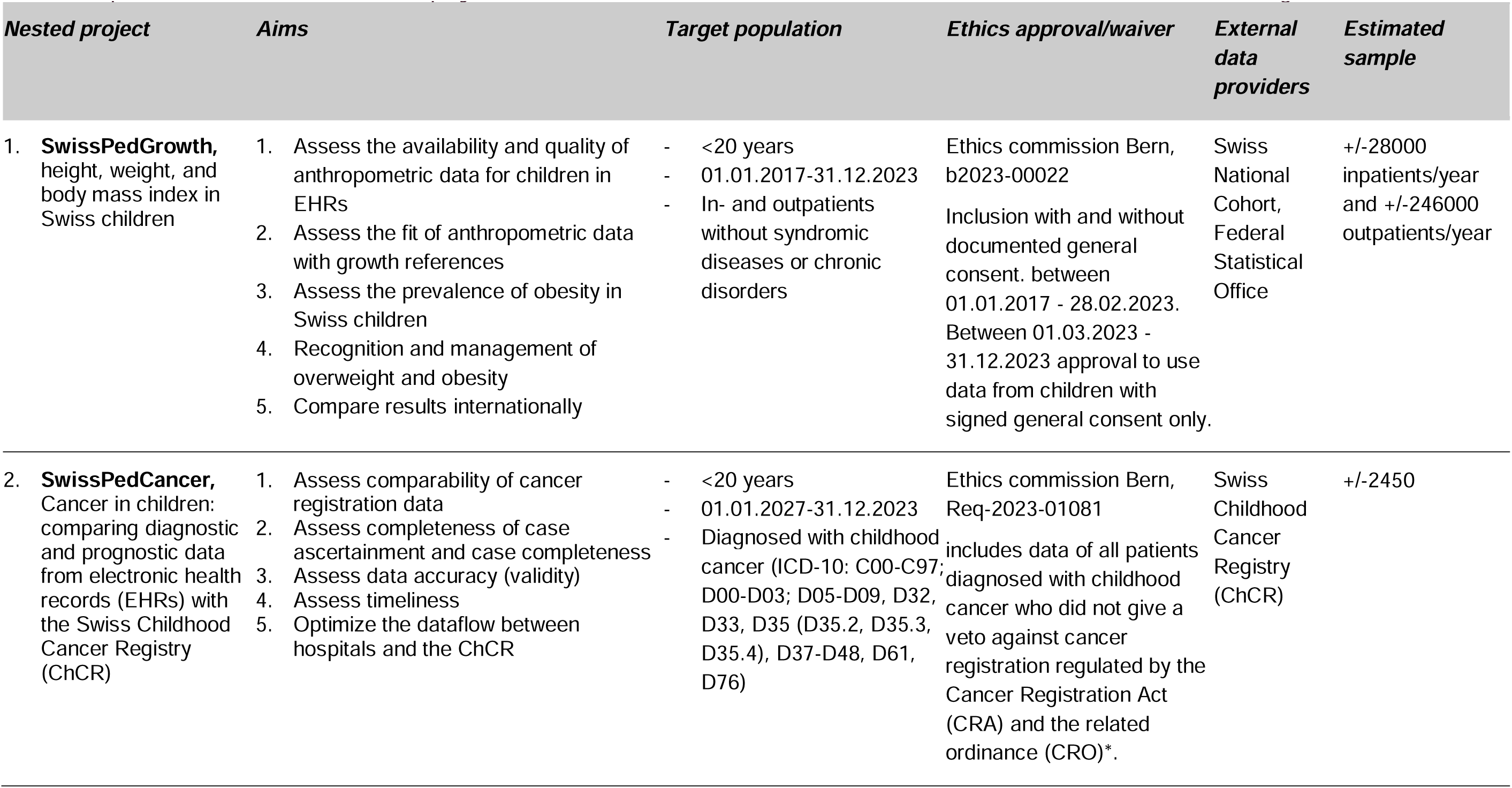
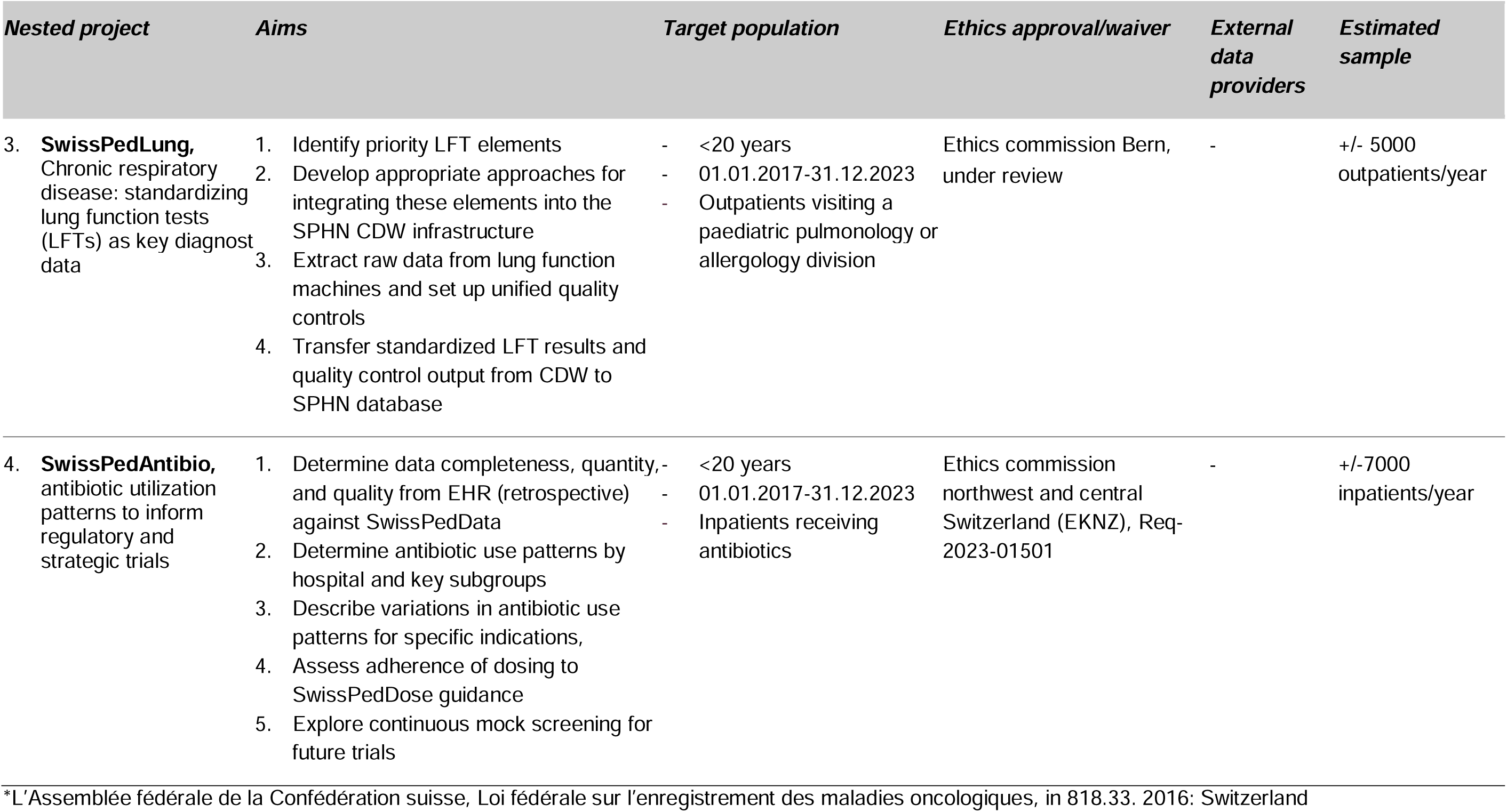
Specifications of the four nested projects within SwissPedHealth: SwissPedGrowth, SwissPedCancer, SwissPedLung, SwissPedAntibio.

All nested projects test availability and quality of information from the electronic health care records, and pilot procedures and data flows. Results of the nested projects will feed back to hospital information systems, and national registries if applicable, to progressively improve data quality. The nested projects are tasked to explore and define strategies to link patient data (such as socioeconomic, nationality, and birth record data) with the Swiss National Cohort^21,22^ and the Federal Statistical Office and compare datasets with external reference standards such as the Swiss Childhood Cancer Registry (https://www.childhoodcancerregistry.ch/).^23,24^ If successful, these linkage strategies will enhance the scalability and interconnectedness of SwissPedHealth, and allow additional assessment of data quality, such as consistency, accuracy, and completeness. Furthermore, the nested projects will partner with international research networks, to identify best practices, compare performance across countries, and enhance the learning of paediatric health systems.

#### The lighthouse project: Development of a multi-omics workflow to discover rare diseases in children with life-threatening phenotypes

The SwissPedHealth lighthouse project focuses on improving diagnosis of rare diseases in children with potentially life-threatening conditions by integrating clinical data with multi-omics data using machine learning algorithms. This project will showcase how the NDS can be enriched with biological data for a specific cohort, in this case children with suspected rare diseases. Rare diseases predominantly affect children, frequently causing premature death or chronic disability.^25^ The discovery of rare diseases has been rapidly increasing in past decades due to decreasing cost coupled with the rising speed of genome sequencing.^26–28^ However, even recent rapid whole-exome or whole-genome sequencing programs in highly selected patients report a diagnostic yield of only 30-50%,^29^ and the challenge has shifted from identifying genetic alterations to defining their functional relevance. Multi-layered omics technologies define relationships between genes, proteins, metabolites, and phenotypic traits, to accelerate identification of the underlying cause of rare clinical presentations.^30^ The lighthouse project will progress through three phases. It builds on our previous pilot testing multi-omics in a defined metabolic disease (phase 1)^31^ to first develop (phase 2), and then prospectively assess (phase 3) a multi-omics workflow designed to identify rare diseases in children with life-threatening conditions (**Fig. 3**).

**Figure 3:**
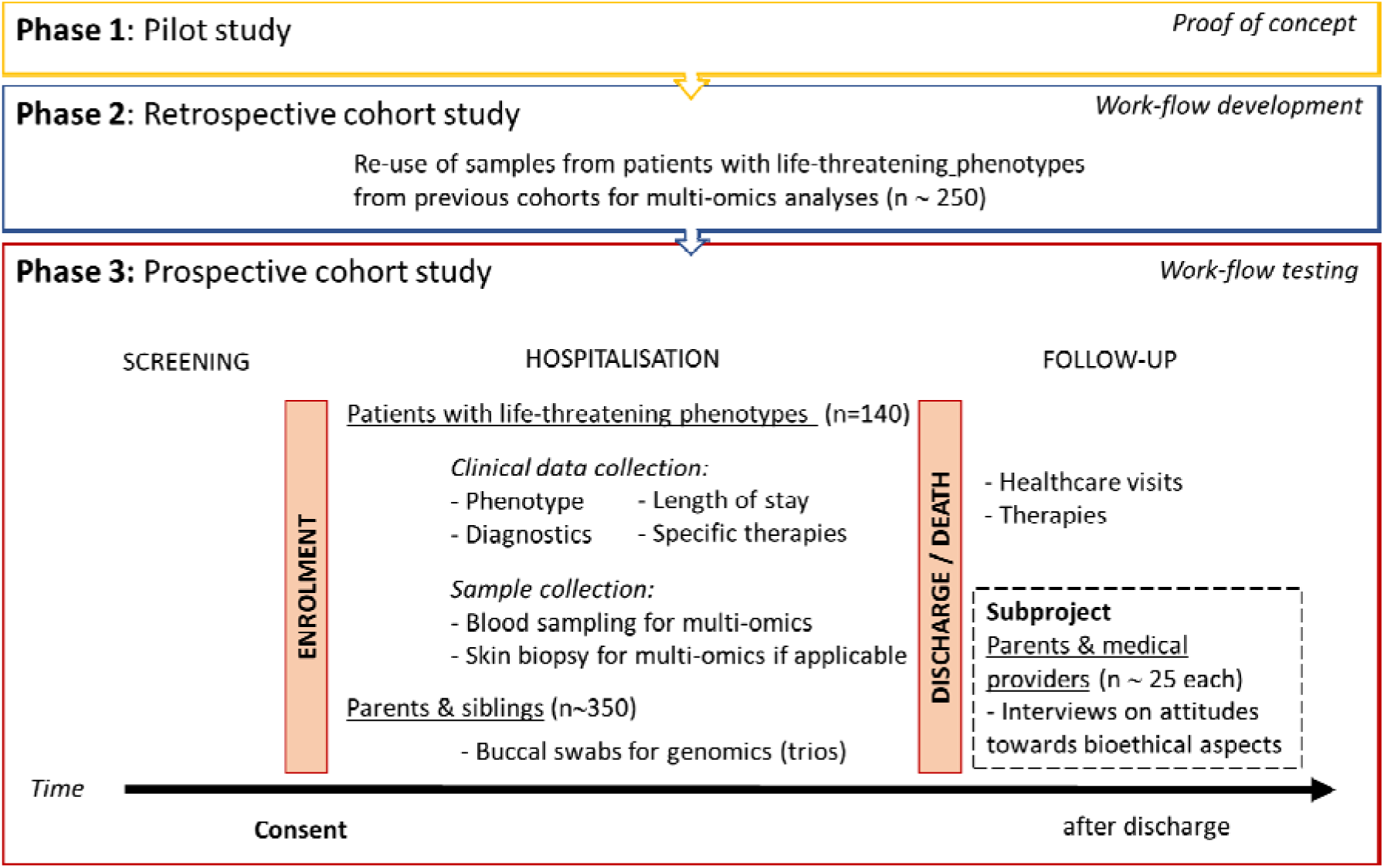
Study design of the SwissPedHealth lighthouse project - Development of a multi-omics workflow to discover rare-diseases in children with life-threatening phenotypes”.

Phase 1 provided a proof of concept for integrative multi-omics analyses in the inborn error of metabolism methylmalonic aciduria (MMA).^31^ In phase 2, we access existing cohorts of children with different life-threatening phenotypes (target N=250), including those who presented with homocystinuria and have a suspected underlying genetic basis, the Swiss Paediatric Sepsis Study (SPSS),^32^ the Genetic Study of Immunodeficiency (GSI),^33^ and Childhood Life-Threatening Infectious Disease Study (EUCLIDS).^34^ We apply an integrated approach of whole-genome sequencing, RNA sequencing,^35–37^ proteomics,^38,39^ and metabolomics,^40^ to identify rare deleterious gene variants and demonstrate their impact at the gene, transcript, and proteome/metabolome level. Samples are processed by the Clinical Genome Analysis Centre, Clinical Proteotype Analysis Centre, and Clinical Metabolomics Analysis Centre. These centres are all part of the Swiss Multi-Omics Centre (http://smoc.ethz.ch/), which is integrated into the PHRT secure computational infrastructure and provides secure data processing and sharing to BioMedIT. Through the lighthouse project, we seek to develop novel bioinformatic pipelines and computational methods using machine learning to integrate multi-omics data with clinical information. This multi-omics workflow will create a diagnostic tool that can be optimized for rapid result generation. In phase 3, we will test and evaluate the workflow developed in phase 2 using a cohort of children prospectively recruited from partner Swiss hospitals (target N=140). Children with potentially life-threatening conditions where a rare disease is suspected are eligible.

Screening occurs through health care staff in the intensive care units and specialist departments, as well as research staff. The eligibility of a patient is assessed by a panel of consortium members including experts in genetics, metabolism, neurology, immunology, and intensive care, before proceeding to seeking informed consent. For each enrolled patient, we collect blood samples (PAX gene for RNA extraction; ethylene-diamine-tetraacetic acid (EDTA) for DNA extraction; serum for proteome/metabolome assessment), and, where feasible, skin biopsies (fibroblasts). In addition, we seek consent to obtain mucosal swabs from parents and siblings of included patients for DNA extraction for trio analyses. Samples are pre-processed at hospitals and sent in batches to the Swiss Multi-Omics Centre for multi-omics data generation. Recruitment for Phase 3 started on September 2022 at the University Children’s Hospital Zurich and will expand to other participating children’s hospitals.

#### Patient and Public Involvement and Bioethics

Patient and public involvement (PPI) represents an integral part of research conduct to increase the quality, meaningfulness, credibility, and impact of clinical research.^41^ Building on existing patient representatives and PPI groups, SwissPedHealth engages with a Patient and Family Advisory Group with diverse members from French and German speaking regions in Switzerland with different roles and contributions (**Fig. 4**). In addition to parents, we will include both adolescent patients and adult survivors of childhood illnesses. The Patient and Family Advisory Group is chaired by two PPI representatives who are part of the Steering Committee. Together with PPI representatives, we develop and test a PPI toolbox to guide future paediatric studies in Switzerland. This toolbox will review PPI involvement and propose practical guidelines that can be used at different time points during the design and conduct of research, as well as the dissemination of study results.

**Figure 4:**
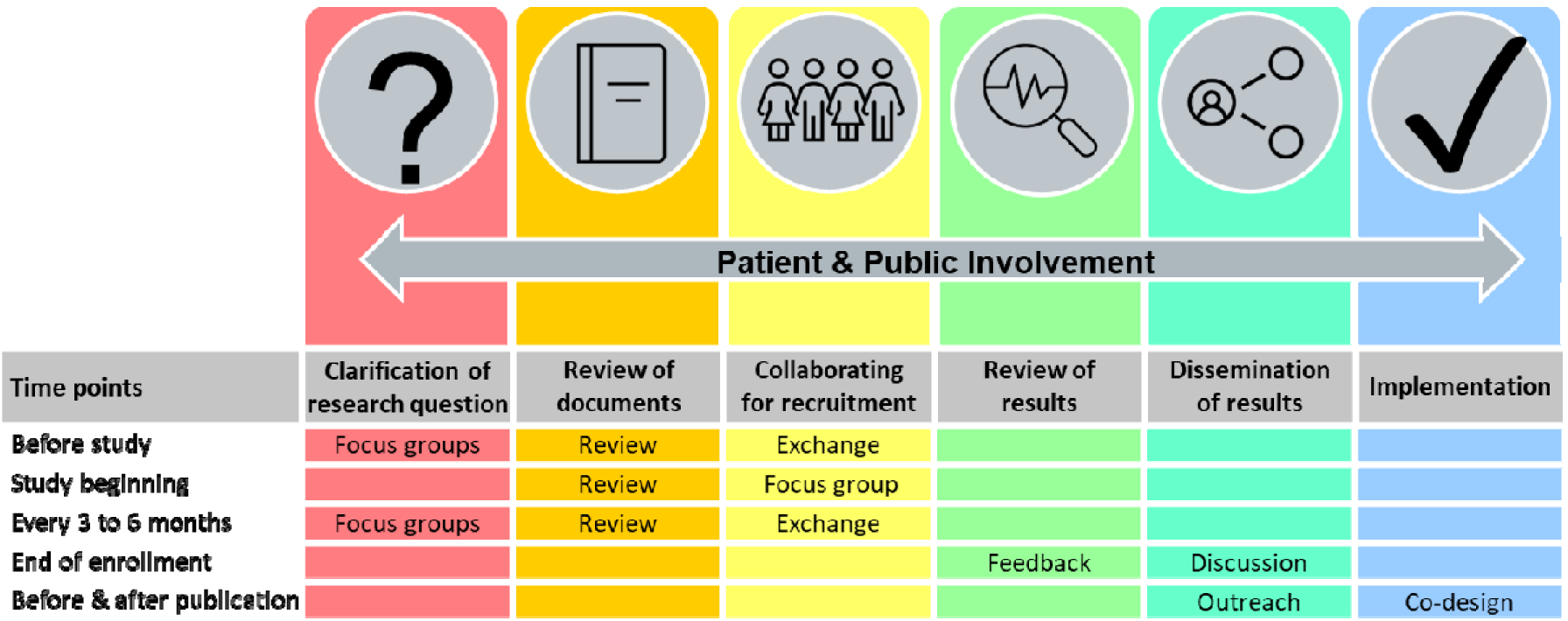
Patient and public involvement activities in SwissPedHealth

Bioethical aspects surrounding perceptions on omics analyses will be explored in depth within a sub-project of the lighthouse project. Given the future goal to translate the current research multi-omics workflow into a clinical standard available to health care providers for patients with suspected genetic rare diseases, it is important to ascertain how this information is viewed by end-users, such as clinicians, patients, and family members. We will interrogate the perspectives of up to 25 medical providers and 25 parents whose children participate in Phase 2 of the lighthouse project by conducting exploratory interviews to determine their attitudes towards a hypothetical offer and return of results and other aspects that will help inform future clinical translation of these technologies.^42^

## ETHICS AND DISSEMINATION

### Ethical approvals

The SwissPedHealth database regulations “Paediatric Personalized Research Network Switzerland (SwissPedHealth)” was approved based on advice of the ethical committee Northwest and Central Switzerland (EKNZ, AO_2022-00018). These regulations have been drawn up in compliance with the applicable standards, considering the Federal Law on Research Involving Human Subjects (Art. 51 Human Research Act), cantonal legislation, and the Federal Data Protection Act. Each nested research or quality improvement project conducted under SwissPedHealth is subject to standard ethical regulatory requirements. Separate ethics approvals and waivers for each nested research or quality improvement project conducted were obtained: SwissPedGrowth (Ethical committee [EC] Bern, b2023-00022); SwissPedCancer (EC Bern, Req-2023-01081); SwissPedLung (EC Bern, under review), SwissPedAntibio (EKNZ, Req-2023-01501), the lighthouse project (EC Zurich, 2022-00351), and the related bioethics interview project (EC ETH Zurich, 2023-N-253).

### Dissemination

The findings of SwissPedHealth, including the lighthouse and nested projects will be published in peer-reviewed journals and scientific conferences, as well as disseminated to patients, families, and the public. Through dedicated PPI focus groups, we will design dissemination to other media, such as the press, radio, television, podcasts, and social media to increase awareness about the NDS, and to promote its use for future research. We will work with families to display understandable and relevant information on our website (https://www.swisspedhealth.ch/).

SwissPedHealth will promote access to analysable datasets and open access publications. Research outcomes will be made publicly available according to the FAIR principles and in line with policies and regulations of the partner institutions and guided by SPHN and PHRT principles.

## DISCUSSION

The paediatric NDS SwissPedHealth digitalizes and harmonizes the use of routine clinical data for clinical research and evidence generation. It will revolutionize paediatric health care in Switzerland in several ways: 1) Personalized care: by harnessing patient data through tailoring health care to individual children’s needs, ensuring the best possible outcomes; 2) Benchmarking: by enabling hospitals to compare their performance with others, identifying areas for improvement; 3) Informing policy decisions: policymakers will have access to data-driven insights, aiding in the development of effective health care policies, and 4) Enabling clinical and therapeutic studies: researchers will be able to use the NDS data to design and conduct studies that lead to more effective treatments for children. SwissPedHealth strives to set up a sustainable framework and infrastructure to conduct Swiss-wide observational and interventional studies, enhancing clinical trial readiness, and to develop a learning health care system driving frontier, innovative, and highly patient-focused research. It seeks to integrate clinical, federal administrative data, and research data across leading Swiss hospitals under joint governance. SwissPedHealth will thereby overcome the challenge of paediatric data being kept in local silos, in non-standardized and non-interoperable forms, highly dispersed across hospitals, and thereby not usable for translational research and clinical trials of sufficient power.

SwissPedHealth is embedded in the SPHN and PHRT Swiss personalized health ecosystem and builds on existing and endurable partnerships with all paediatric teaching hospitals centred around SwissPedNet. It will implement a harmonized paediatric health dataset across Switzerland, which is scalable to additional datasets and sites, and serves as a high-visibility example of how this infrastructure can be enriched with high-density data to deliver cutting-edge research on personalized health. SwissPedHealth also builds on previous successful national multi-centre initiatives such as SwissPedData, the Swiss Pharmacokinetics Clinical Data Warehouse project (SwissPKcdw),^43^ the Swiss Paediatric Sepsis Study,^44,45^ and the Genetic Study of Immunodeficiency (GSI).^46–49^ The broad scope of SwissPedHealth also favours potential synergies with other current SPHN and PHRT funded projects (https://sphn.ch/network/projects/, such as LUCID, IICU, SPHN-SPAC, SwissPedDW, SwissPedHealth-PREPP, ADONIS, INFRA).

Internationally, SwissPedHealth shares goals and strategies of several initiatives, such as the European Children’s Hospital Organization (ECHO, https://www.echohospitals.org/), and the US based paediatric clinical research network (PEDSnet, https://pedsnet.org/) which use a common interoperable data platform to optimize the use of electronic health records.

There are several challenges to be overcome in SwissPedHealth. Regulatory processes such as legal agreements and multi-centre ethic proposals that need to be signed by several hospitals and institutions are cumbersome, despite standardized templates. The design of RDF concepts requires multi-disciplinary expertise in content aspects, like medicine and biology, as well as RDF, semantic and technical knowledge. Data harmonization efforts at local CDWs are time consuming and require both technically and clinically skilled staff, and adequate local resources. Source systems are different in every hospital, making efforts in one site often not applicable or directly comparable to other sites. Central coordination efforts are crucial to create synergies whenever possible. There is heterogeneity in data availability between CDWs. Thus, data gaps are expected for the nested and lighthouse projects of SwissPedHealth. Regulatory barriers need to be overcome to integrate patient data from sources other than hospitals such as registries and federal offices which have their own regulations and data structure. Data harmonization efforts have primarily focused on University Hospitals; however, additional efforts are necessary to extend coverage to non-university clinics and primary care, thereby including a broader segment of the Swiss child population. Future endeavours will facilitate and explore the data interoperability with existing international data models such as Observational Medical Outcomes Partnership (OMOP) Common Data Model.^15^

### Outlook

We will set up a sustainable framework and infrastructure to conduct Swiss-wide observational and interventional studies, enhancing clinical trial readiness, and to develop a learning health care system driving frontier, innovative, and highly patient-focused research. In the future, the experience gained and the infrastructure and procedures developed within SwissPedHealth are expected to facilitate integration of other Swiss paediatric hospitals, and linkage with different databases, such as Swiss paediatric national registries,^23,44,50–56^ as well as exchange with international consortia.

The vision of SwissPedHealth is a future where routine paediatric data from hospitals, and ideally also primary care providers, other research institutions, biobanks, and registries, can be securely used to sustain a learning health care system and for research to improve and personalize health care of children. This framework within a Swiss personalized health ecosystem is only possible through joint, and highly coordinated efforts from clinicians, engineers, scientists, researchers, legal and regulatory experts, and patients and families.

## Data Availability

Research outcomes will be made publicly available according to the FAIR principles and in line with policies and regulations of the partner institutions and guided by Swiss Personalized Health Network (SPHN) and Personalized Health and Related Technologies (PHRT) principles.

https://www.swisspedhealth.ch/

## Acknowledgements

We would like to thank all the patients and families who have been providing advice on SwissPedHealth and its projects, as well as the clinical and research teams at the participating institutions.

## SwissPedHealth consortium

- Andrea Agostini, Department of Computer Science, Institute for Machine Learning, ETH Zurich, Zurich, Switzerland
- Anita Rauch, Institute of Medical Genetics, University of Zurich, Zurich, Switzerland
- Anna Hartung, Inselspital, Bern University Hospital, University of Bern, Switzerland
- Audrey van Drogen, PHRT Swiss Multi-Omics Centre (SMOC), ETH Zurich, Zurich, Switzerland & Institute of Translational Medicine (ITM), Department of Health Sciences and Technology (D-HEST), ETH Zurich, Zurich, Switzerland
- Aurélie Martin Necker, Patient and Family Advisory Committee, SwissPedHealth
- Ben D. Spycher, Institute of Social and Preventive Medicine (ISPM), University of Bern, Bern, Switzerland
- Christian Kahlert, Ostschweizer Kinderspital, St Gallen, Switzerland
- Christopher B. Forrest, Center for Applied Clinical Research, Children’s Hospital of Philadelphia, Philadelphia, USA
- Claudia E. Kuehni, Institute of Social and Preventive Medicine (ISPM), University of Bern, Bern, Switzerland & Division of Paediatric Respiratory Medicine and Allergology, Children’s University Hospital, Inselspital, University of Bern, Bern, Switzerland
- Cornelia Hagman, Department of Intensive Care and Neonatology and Children’s Research Centre, University Children’s Hospital Zurich, University of Zurich, Zurich, Switzerland
- D Sean Froese, Division of Metabolism and Children’s Research Centre, University Children’s Hospital Zurich, University of Zurich, Zurich, Switzerland
- Daphné Chopard, Department of Computer Science, Institute for Machine Learning, ETH Zurich, Zurich, Switzerland & Department of Intensive Care and Neonatology and Children’s Research Centre, University Children’s Hospital Zurich, University of Zurich, Zurich, Switzerland
- Dylan Lawless, School of Life Sciences, Ecole Polytechnique Fédérale de Lausanne, Lausanne, Switzerland & Department of Intensive Care and Neonatology and Children’s Research Centre, University Children’s Hospital Zurich, University of Zurich, Zurich, Switzerland
- Effy Vayena, Department of Health Sciences and Technology, Institute of Translational Medicine, ETH Zurich, Zurich, Switzerland
- Eirini I. Petrou, Department of Health Sciences and Technology, Institute of Translational Medicine, ETH Zurich, Zurich, Switzerland
- Emanuele Palumbo, Department of Computer Science, Institute for Machine Learning, ETH Zurich, Zurich, Switzerland
- Eric Giannoni, Clinic of Neonatology, Lausanne University Hospital and University of Lausanne, Lausanne, Switzerland
- Fabiën N. Belle, Institute of Social and Preventive Medicine (ISPM), University of Bern, Bern, Switzerland
- Ioannis Xenarios, PHRT Swiss Multi-Omics Centre (SMOC), EPFL, Lausanne, Switzerland & Department of Computational Biology, University of Lausanne, Lausanne 1015, Switzerland & Health 2030 Genome Center, Foundation Campus Biotech, Geneva, Switzerland.
- Jacques Fellay, School of Life Sciences, Ecole Polytechnique Fédérale de Lausanne, Lausanne, Switzerland & Biomedical Data Science Center, Lausanne University Hospital and University of Lausanne, Lausanne, Switzerland
- Jana Pachlopnik Schmid, Division of Immunology and Children’s Research Centre, University Children’s Hospital Zurich, University of Zurich, Zurich, Switzerland
- Julia A. Bielicki, Paediatric Research Center, University l Children’s Hospital Basel (UKBB), University of Basel, Basel, Switzerland & Centre for Neonatal and Paediatric Infection, St George’s, University of London, London, UK
- Julia E. Vogt, Department of Computer Science, Institute for Machine Learning, ETH Zurich, Zurich, Switzerland
- Kathrin Hofmann, Patient and Family Advisory Committee, SwissPedHealth
- Katrin Männik, PHRT Swiss Multi-Omics Centre (SMOC), EPFL, Lausanne, Switzerland & Center for Integrative Genomics, University of Lausanne, Lausanne, Switzerland & Health 2030 Genome Center, Foundation Campus Biotech, Geneva, Switzerland.
- Keith Harshman PHRT Swiss Multi-Omics Centre (SMOC), EPFL, Lausanne, Switzerland & Health 2030 Genome Center, Foundation Campus Biotech, Geneva, Switzerland
- Kelly Ormond; Department of Health Sciences and Technology, Institute of Translational Medicine, ETH Zurich, Zurich, Switzerland & Department of Genetics, Stanford University School of Medicine, Stanford, California, US
- Klara Posfay-Barbe, Hôpitaux Universitaires de Genève, Geneva, Switzerland
- Léa Ho Dac, Division of Paediatric Respiratory Medicine and Allergology, Department of Paediatrics, Inselspital, Bern University Hospital, University of Bern, Switzerland
- Lorenz M. Leuenberger, Institute of Social and Preventive Medicine (ISPM), University of Bern, Bern, Switzerland
- Luregn J Schlapbach, Department of Intensive Care and Neonatology and Children’s Research Centre, University Children’s Hospital Zurich, University of Zurich, Zurich, Switzerland & Child Health Research Centre, The University of Queensland, Brisbane, Australia
- Manon Jaboyedoff, Pediatric Infectious Diseases and Vaccinology Unit, Service of Pediatrics, Department Mother-Woman-Child, Lausanne University Hospital and University of Lausanne, Lausanne, Switzerland
- Mariam Ait Oumelloul, School of Life Sciences, Ecole Polytechnique Fédérale de Lausanne, Lausanne, Switzerland
- Martin Stocker, Luzerner Kantonsspital, Luzern, Switzerland
- Matthias R Baumgartner, Division of Metabolism and Children’s Research Centre, University Children’s Hospital Zurich, University of Zurich, Zurich, Switzerland
- Nicola Zamboni, PHRT Swiss Multi-Omics Centre (SMOC), ETH Zurich, Zurich, & Institute of Molecular Systems Biology, ETH Zurich, Zurich, Switzerland
- Patrick G. A. Pedrioli, PHRT Swiss Multi-Omics Centre (SMOC), ETH Zurich, Zurich, Switzerland & Institute of Translational Medicine (ITM), Department of Health Sciences and Technology (D-HEST), ETH Zurich, Zurich, Switzerland & Swiss Institute of Bioinformatics, Lausanne, Switzerland & Department of Biology, Institute of Molecular Systems Biology, Swiss Federal Institute of Technology/ETH Zürich, Zurich, Switzerland
- Philipp Latzin, Division of Paediatric Respiratory Medicine and Allergology, Department of Paediatrics, Inselspital, Bern University Hospital, University of Bern, Switzerland
- Rebeca Mozun, Department of Intensive Care and Neonatology and Children’s Research Centre, University Children’s Hospital Zurich, University of Zurich, Zurich, Switzerland
- Roger Lauener, Ostschweizer Kinderspital, St Gallen, Switzerland
- Sandra Goetze, PHRT Swiss Multi-Omics Centre (SMOC), ETH Zurich, Zurich, Switzerland & Institute of Translational Medicine (ITM), Department of Health Sciences and Technology (D-HEST), ETH Zurich, Zurich, Switzerland.
- Seraina Prader, Division of Immunology and Children’s Research Centre, University Children’s Hospital Zurich,
- Simon Boutry, School of Life Sciences, Ecole Polytechnique Fédérale de Lausanne, Lausanne, Switzerland
- Sven Schulzke, Department of Neonatology, University Children’s Hospital Basel (UKBB), University of Basel, Basel, Switzerland
- Tatjana Welzel, Paediatric Research Center, University Children’s Hospital Basel (UKBB), University of Basel, Basel, Switzerland
- Thomas M. Sutter, Department of Computer Science, Institute for Machine Learning, ETH Zurich, Zurich, Switzerland
- Varvara Dimopoulou, Clinic of Neonatology, Lausanne University Hospital and University of Lausanne, Lausanne, Switzerland
- Vito RT Zanotelli, Division of Metabolism and Children’s Research Centre, University Children’s Hospital Zurich, University of Zurich, Zurich, Switzerland
- Xenia Bovermann, Division of Paediatric Respiratory Medicine and Allergology, Department of Paediatrics, Inselspital, Bern University Hospital, University of Bern, Switzerland
- Yara Shoman, Institute of Social and Preventive Medicine (ISPM), University of Bern, Bern, Switzerland Division of Immunology and Children’s Research Centre, University Children’s Hospital Zurich, Switzerland

